# Assessment of Model Estimated and Directly Observed Weather Data for Etiological Prediction of Diarrhea

**DOI:** 10.1101/2023.10.12.23296959

**Authors:** Ben J. Brintz, Josh M. Colston, Sharia M. Ahmed, Dennis L. Chao, Ben Zaitschik, Daniel T. Leung

## Abstract

Recent advances in clinical prediction for diarrheal etiology in low- and middle-income countries have revealed that addition of weather data improves predictive performance. However, the optimal source of weather data remains unclear. We aim to compare model estimated satellite- and ground-based observational data with weather station directly-observed data for diarrheal prediction. We used clinical and etiological data from a large multi-center study of children with diarrhea to compare these methods. We show that the two sources of weather conditions perform similarly in most locations. We conclude that while model estimated data is a viable, scalable tool for public health interventions and disease prediction, directly observed weather station data approximates the modeled data, and given its ease of access, is likely adequate for prediction of diarrheal etiology in children in low- and middle-income countries.

---

Infectious diarrhea is a significant public health concern, particularly in low resource settings where access to clean water and sanitation is limited. The incidence of infectious diarrhea is influenced by a variety of factors, including seasonality and weather conditions (Chao et al. (2019)). Previous studies have shown that weather conditions, such as precipitation, are strongly associated with the incidence of infectious diarrhea in low and middle-income countrys (LMICs) settings (Colston et al. (2022), Horn et al. (2018)). Weather conditions in the months prior to illness have also been found to be predictive of the incidence of specific causes of diarrheal illness, such as rotavirus (Colston et al. (2018)) and Shigella (Badr et al. (2023)). In addition, we have previously shown that directly observed (DO) data from weather stations accessed through the Global Surface Summary of the Day online repository can be used to derive the local season (Brintz et al. (2020)) or a moving average of recent weather conditions (Brintz et al. (2021)) and incorporated with clinical variables to improve prediction of the etiology of infectious diarrhea in children living in LMICs. Such etiological prediction models can then be adapted to clinical decision-support tools for improved stewardship of antimicrobial and diagnostic use (Nelson et al. (2022)). Gridded meteorological estimates from models using satellite- and ground-based observational data, such as that obtained from the Global Land Data Assimilation System (GLDAS), enable access to granular temperature and precipitation data over time and space that are more consistent than the DO weather station data, though requires more processing. In this analysis, we use data from a large multi-center case-control study of children with diarrhea to assess and compare the performance of the model estimated (ME) data of GLDAS to the DO weather station data of the Global Historical Climatology Network daily (GHCNd) when making predictions of diarrheal episode etiology.

We tested the predictive performance of the weather data applied to the Global Enteric Multicenter (GEMS) Study, which we previously used to derive and test prediction models for a viral only etiology vs. other etiologies of diarrhea (Brintz et al. (2020), Brintz et al. (2021), Garbern et al. (2022)). Briefly, the GEMS was an observational case-control study conducted between 2007 and 2011 at healthcare facilities in 7 countries, in which 9439 children with moderate-to-severe diarrhea were enrolled at local health care centers along with one to three matched community control-children. A fecal sample was taken from each child at enrollment to identify enteropathogens and clinical information was collected. We used the quantitative real-time PCR-based (qPCR) attribution models developed by Liu et al. (2016) in order to best characterize the cause of diarrhea. We defined viral etiology as a diarrhea episode with at least one viral pathogen with an episode-specific attributable fraction (AFe 0.5) and no bacterial or parasitic pathogens with an episode-specific attributable fraction. Prediction of viral attribution is clinically meaningful since it indicates that a patient would not benefit from antibiotic therapy.

We used daily aggregates of gridded meteorological estimates extracted from GLDAS, which includes 3-hourly weather information on a 25 × 25 kilometer grid, based on the locations of GEMS hospital for each country (Rodell et al. (2004), Colston et al. (2022)). The temperature is extracted as minimum and maximum daily temperature. We averaged the daily minimum and maximum temperature in order to calculate an average daily temperature. Although, the average of the min and max is not necessarily equivalent to the daily average, prior literature has shown agreement between this approach and various other approaches used to get a daily temperature summary as well as ground-based measures (Weiss and Hays (2005)). We additionally extracted the total daily precipitation in millimeters.

We used the GHCNd to obtain DO average daily temperature and daily total precipitation. The GHCNd combines daily observations from over 30 different sources of climate observations, and undergoes a quality assurance process approximately weekly (Menne et al. (2012)). We utilized the **readr** package in R to directly download station data from GHCN daily during the study period. For each GEMS study site, we selected the closest weather station, based on haversine distance, that contained data during the GEMS years. For each location, missing weather data was completed using the most recent non-missing weather data prior to it from that same station.

We used logistic regression as the main model type for assessing the predictive performance of weather variables on viral versus other etiology. We fit a separate model for each of the following moving windows of exposure in which the interval contains the days prior *t*_−*d*_ to the current day *t*_0_ included in the average:

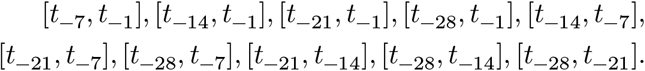

Each model contained three exposure covariates: 1) temperature and 2) precipitation and 3) their interaction.

With this approach, we assess whether certain intervals of weather conditions prior to the presentation of the patient are more predictive of viral etiology than others, including some intervals which don’t start immediately prior to the day of presentation. Finally, we assess the performance of cross-validated predictions by site. We order them by distance to the reporting weather station to assess whether proximity may have been a factor in predictive performance.

We assess the predictive performance of the model fits with various performance metrics using repeated cross-validation. We generated 500 random 80% training and 20% testing splits and compared model performance between GLDAS data and station data using the models with covariates calculated using the intervals described above. Within each iteration, we calculated the area under the receiver operating characteristic curve (AUC), the calibration intercept and slope of the model for assessing weak-calibration, the area under the precision-recall curve (PRAUC), and we used bootstrapping to compare the ROC curves between models (Van Calster et al. (2019), DeLong, DeLong, and Clarke-Pearson (1988)). The PRAUC, though less common than the AUC, can be more pertinent when assessing performance of predictive models for outcomes with imbalanced classes due to its emphasis on the performance when the ground truth is the positive class. In this case, viral etiologies account for approximately 1/3 of the outcomes.

We found that when we plotted the ME and DO temperature and the precipitation in time series by site, they were very similar. However, the Modeled estimates outperform the directly observed data in both AUC and PRAUC at each interval explored with the best AUC from each source being 0.66 and 0.64, and the best PRAUC from each source being 0.47 and 0.44, respectively (Figure 1). The best performing models tend to include weather terms using moving windows which include the most recent days from the incident infection.

**Figure 1:**
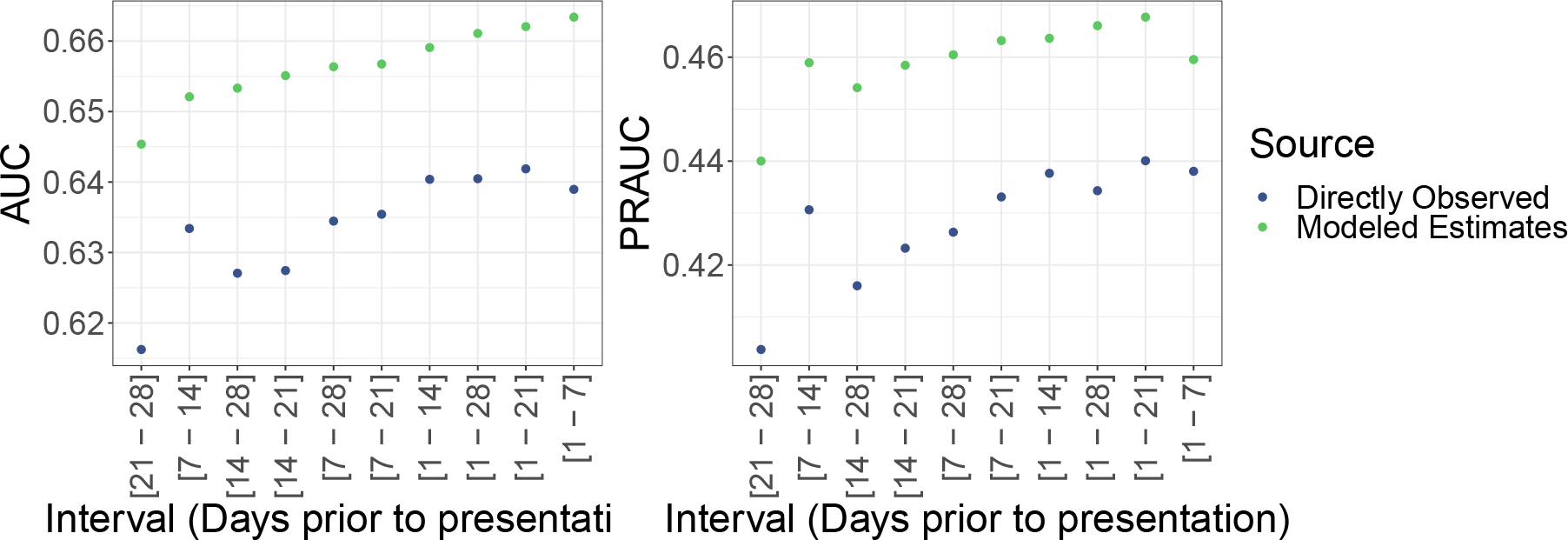
Overall cross-validated AUC and PRAUC for model estimated and directly observed weather data using various intervals to average over prior to the day of the prediction.

When predictions from cross-validation are broken down by site, the superior predictive performance using ME over DO data is limited to The Gambia with a difference in average AUC of 0.102, or about 10%. Notably, the Gambia’s clinical study site is 120 km away from the closest weather station, the fourth farthest site from its weather station. The next largest difference in average AUC between data sources by site in the [1-7] interval was 0.038 in Mali. When we use bootstrapping within each iteration of cross-validation to compare AUC’s between models driven by the two sources of data using the 1-7 interval for all sites, we find that the median p-value in cross-validation is 0.025. This suggests that there is evidence of a difference between models fit using the two sources of data; however, when we conduct the same bootstrapping test and compare the two sources of data within sites, Gambia has a median p-value of 0.055, while the other sites have median p-values of 0.291 and above. We additionally found that all models satisfied the criteria for weak calibration, i.e., the estimates for the calibration intercept were close to 0 with 95% CIs that contain 0 and the calibration slopes were close to 1 with 95% CIs that contain 1.

Clinicians in low-resource settings are often required to make clinical decisions of infectious syndromes without information from laboratory diagnostics. Traditional clinical prediction rules typically focus on data from the patient at presentation, but more recently, the use of location-specific (or patient-extrinsic) data sources such as climate have shown to improve the performance of prediction rules over clinical factors alone (Brintz et al. (2020), Brintz et al. (2021)). The optimal method for estimating climate and weather data for incorporation into clinical prediction rules has not been assessed. We demonstrate that overall, while DO weather station data vary in availability, they require less technical expertise, and ultimately show similar predictive performance as the ME of weather based on satellite- and ground-based observation, for prediction of etiology of diarrhea in pediatric patients.

In both methods, though most prominently with ME weather, we found that the top 4 models by AUC all included temperature and precipitation data with the shortest lag length, (i.e., included the prior days’ weather data in the aggregate predictors), consistent with the known short incubation periods of most enteropathogens, and previous findings observed with individual organisms (Colston et al. (2022)). The top 3 models by PRAUC also included the shortest lag.

We note some discrepancies in data preparation between the two sources of data. First, the DO data are less complete than the ME, e.g., precipitation missing in periods of time where there is no precipitation. In contrast, by its nature, the model estimates have no missing data. However, when using this estimated data, advanced data storage and data cleaning will likely be required for any given application due to its granularity in raw form. Although we find evidence of an overall difference between predictive performance, this difference can be attributed to differences at a single site (The Gambia). We did not find a clear linear trend in performance difference when looking at distance from closest weather station. Further explorations should be made to assess if the best performing lags from the two sources of weather data apply to other prediction outcomes, and whether our findings can be generalized to other climate factors such as humidity or environmental factors such as air pollution.

In conclusion, we found that GHCNd’s DO weather station-derived data approximates GLDAS’s ME meteorologic data, and is likely adequate, for prediction of diarrheal etiology in children in LMICs.

## Data Availability

All data produced in the present study are available upon reasonable request to the authors

https://www.ncei.noaa.gov/products/land-based-station/global-historical-climatology-network-daily

